# Third BNT162b2 vaccination neutralization of SARS-CoV-2 Omicron infection

**DOI:** 10.1101/2021.12.13.21267670

**Authors:** Ital Nemet, Limor Kliker, Yaniv Lustig, Neta S. Zuckerman, Oran Erster, Carmit Cohen, Yitshak Kreiss, Sharon Alroy-Preis, Gili Regev-Yochay, Ella Mendelson, Michal Mandelboim

## Abstract

Using isolates of SARS-CoV-2 WT, Beta, Delta and most importantly Omicron we studied the capability of the BNT162b2 vaccine given in two or three doses to neutralize major SARS-CoV-2 variants of concern (VOC).

We demonstrate low neutralization efficiency against delta and wild-type for vaccines with more than 5 months following the second BNT162b2 dose, with no neutralization efficiency against Omicron. We demonstrate the importance of a third dose, by showing a 100-fold increase in neutralization efficiency of Omicron following a third dose, with a 4-fold reduced neutralization compared to that against the Delta VOC. The durability of the effect of the third dose is yet to be determined.

---

On November 26, 2021, the World Health Organization (WHO) named the B.1.1.529 COVID-19 variant, first detected in South Africa, as the Omicron variant of concern (VOC) (1). By 29 November, 2021, three days after the announcement by WHO, cases of VOC Omicron have already been detected in many other countries.

Whether the BNT162b2 vaccine, that was previously reported to have 95% efficacy against coronavirus disease 2019 (Covid-19) (2,3) will effectively neutralize Omicron infection is unknown. Here, we compared neutralization of Omicron infected cells by sera of 2-dose vaccinated vs. 3-dose vaccinated individuals.

A micro-neutralization assay with Wild Type, Beta, Delta and Omicron isolates was performed using 20 serum samples obtained from two groups of health care workers (HCW): (1) vaccinated by two doses of BNT162b2, five to six months after the second dose and (2) HCW vaccinated by three doses, one month after the third dose (Table S1). Third vaccination lead to a better neutralization of all viruses (Fig. 1). The Geometric Mean Titers (GMT) of WT, Beta, Delta and Omicron were 16.56, 1.27, 8 and 1.11, respectively, after the second vaccination and 891.4, 152.2 430.5 and 107.6, respectively after the third vaccination (Fig. 1). A significant reduction in BNT162b2 neutralization efficiency against all Variants of Concern (VOC): Beta, Delta and Omicron, as compared to the WT virus, was observed when comparing two or three vaccine doses (Fig. 1B and D). Interestingly, the reduction in neutralization efficiency against the beta and Omicron variants as compared to the WT virus, was similar, both after the second (Fig. 1 B) and after the third vaccination (Fig. 1 D). Importantly, third dose of BNT162b2 vaccine, efficiently neutralized Omicron infection (GMT 1.1 following second dose vs. 107.6 following third dose, Fig. 1 A and C, respectively).

**Figure 1:**
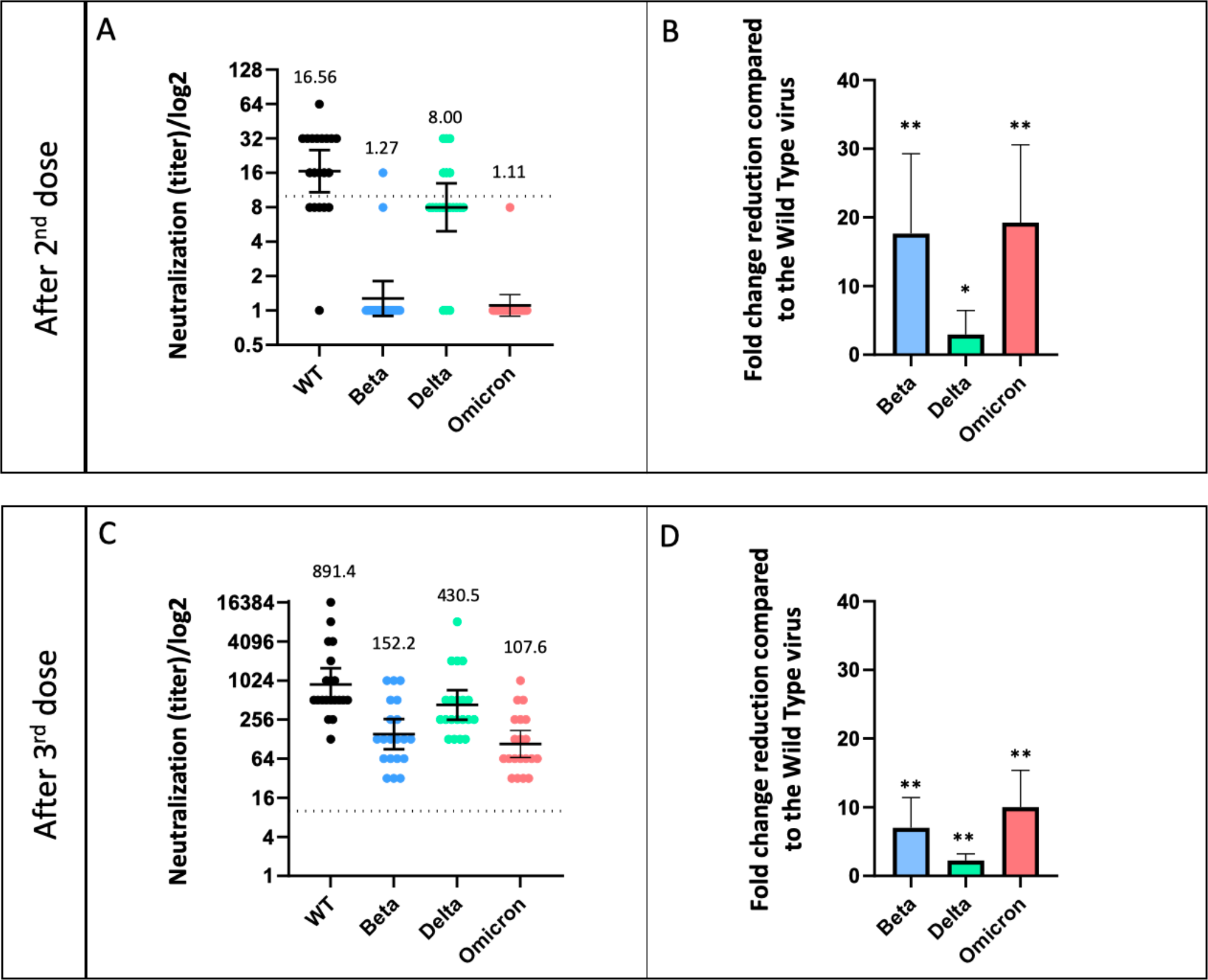
Neutralization efficiency: Sera obtained from 20 HCW vaccinated with two (A-B) and three doses (C-D), were tested by micro-neutralization against the various viruses (presented on the X axis). Fold change reduction as compared to the WT virus is shown after second (B) and third dose (C). The dashed line indicates cut off titer. Geometric mean titers (GMT) with 95% CI are presented as well as the GMT value for each time point.

We analyzed the neutralization efficiency against wild-type, Beta, Delta and Omicron VOC. Limitations of the study include the small cohort tested and this test being only an in-vitro assay. Yet, we demonstrate low neutralization efficiency against delta and wild-type for vaccines with more than 5 months following the second BNT162b2 dose, with no neutralization efficiency against Omicron. We demonstrate the importance of a third dose, by showing a 100-fold increase in neutralization efficiency of Omicron following a third dose, with a 4-fold reduced neutralization compared to that against the Delta VOC. The durability of the effect of the third dose is yet to be determined.

## Data Availability

All data produced in the present study are available upon reasonable request to the authors

