## Supplementary Information for "Third BNT162b2 vaccination neutralization of SARS-CoV-2 Omicron infection"

This appendix has been provided by the authors to give readers additional information about their work.

**Table of content**

Supplemental Materials and Methods 3

• Cohort 3

• Ethical statement 3

• Viral isolation of the Wild Type, Beta, Delta and Omicron variants 3

• Viral titration 4

• SARS-CoV-2 micro-neutralization assay 4

• Specific detection of Omicron SARS-CoV-2 Variant By RT-qPCR differential assays 4

• Statistical Methods 5

• Acknowledgments 5

• Bibliography 6

Table S1 (Demographics of study participants included in this report) 6

### Supplementary information:

#### Methods:

#### Cohort

Sheba Medical Centre is the largest tertiary medical center in Israel, with over 14,000 HCW. Two groups were randomly picked from the Sheba HCW COVID cohort, if they meet the inclusion criteria: (1) Not previously infected by SARS-CoV-2 as determined by PCR, pre-vaccine serology testing, or anti-N IgG on follow-up. (2) Received the first two BNT162b2 doses at least 5 months previously. Samples of 20 HCW that received the second BNT162b2 vaccine dose but not the third dose and another 20 samples from HCW who received the third BNT162b2 vaccine dose one month previously. (Table S1).

#### Ethical statement

The protocol was approved by the Institutional review board of the Sheba Medical Center. Written informed consent was obtained from all participants.

#### Viral isolation of the Wild Type, Beta, Delta and Omicron variants

Using sequencing we identified 4 nasopharyngeal samples from SARS-CoV-2 positive individuals which contained the Wild Type sub lineage B.1.1.50 (hCoV-19/Israel/CVL-45526-ngs/2020), Beta, B.1.351, (hCoV-19/Israel/CVL-2557-ngs/2020), Delta, B.1.617.2 (hCoV-19/Israel/CVL-12804/2021) and B.1.1.529, Omicron (hCoV-19/Israel/SMC-7023409/2021) variants. Confluent VERO-E6 cells were incubated for one hour at $33^{0}C$ with 300 µl of nasopharyngeal samples followed by addition of 5 ml 2% FCS MEM-EAGLE medium. Upon CPE detection, supernatants were aliquoted and stored at -$80^{0}C$.

**Viral titration**

In order to calibrate and determine the 50% endpoint titer (TCID50) of each variant VERO-E6 cells at concentration of $20*10^3$/well, were seeded in three sterile 96-wells plates with 10% FCS MEM-EAGLE medium, and stored at $37^oC$ for 24 hours. Ten-fold serial dilutions of each variant were prepared using 2% FCS MEM-EAGLE medium and incubated for five days with the VERO-E6 cells. Following Gentian Violet staining TCID50 of each variant was calculated using the Spearman-Karber method.

**SARS-CoV-2 micro-neutralization assay**

VERO-E6 cells at concentration of 20*103/well were seeded in sterile 96-wells plates with 10% FCS MEM-EAGLE medium, and stored at $37^oC$ for 24 hours. One hundred TCID50 of Wild Type, Beta, Delta and Omicron SARS-CoV-2 isolates were incubated with inactivated sera diluted 1:10 to 1:16,384 in 96 well plates for 60 minutes at 33ºC. Virus-serum mixtures were added to the Vero E-6 cells and incubated for five days at 33ºC after which Gentian violet staining (1%) was used to stain and fix the cell culture layer. Neutralizing dilution of each serum sample was determined by identifying the well with the highest serum dilution without observable cytopathic effect. A dilution equal to 1:10 or above was considered neutralizing.

**Specific detection of Omicron SARS-CoV-2 Variant By RT-qPCR differential assays**

Four RT-qPCR assays that enable rapid identification of the newly emerging SARS-COV-2 Omicron (B.1.1.529) variant of concern (1). The assays target Omicron characteristic mutations in the nsp6 (Orf1a), spike and nucleocapsid genes. We demonstrate that the assays are straightforward to assemble and perform, are amendable for multiplexing, and may be used as a reliable first-line tool to identify B.1.1.529 suspected samples.

**Statistical Methods**

Neutralizing antibodies and Geometric Mean Titers (GMT) with confidence interval (CI) of 95% were performed using GraphPad Prism 5.0 (GraphPad Software, Inc., San Diego, CA).

Wilcoxon matched-pairs signed-rank test was performed to detect significant differences in neutralizing titers between the Wild Type and the other variants

**Acknowledgments**

We wish to acknowledge the contribution of the following groups: 1. members of the SARS CoV-2 sequencing team from Shamir Medical Center, Israel, headed by Dr. Adina Bar-Chaim, who provided the full sequence of the B.1.1529 (Omicron) isolate used in this study. 2. Israel Defense Forces (IDF) units which helped to track down cases carrying the B.1.1.529 (Omicron) variant and to obtain clinical samples for diagnosis and culturing. 3.Members of the Ministry of Health team which coordinates the National Sequencing Operation from sample collection to data analysis. 4. Dr Ofra Havkin, head of the National Committee for SARS CoV-2 genomic sequencing. 5. Donors of the serum samples involved in this study. We wish to thanks all these people for allowing us to conduct this important study.

**Supplementary Table 1: Demographics of study participants included in this report**

| | Two vaccinations | Three vaccinations |
|---|---|---|
| Number of participants | 20 | 20 |
| Age (Median) | 24-74 (54.725) | 24-56 (38.74) |
| Average age | 53.3935 | 37.77526 |
| Males | 6 | 1 |
| Females | 14 | 19 |
| Days post vaccination (Avg) | 154-176 (165.6) | 15-31 (25) |
